# Combined Effects of Botulinum Toxin Therapy and Splint Therapy on Upper Limb Spasticity in Chronic Stroke Patients: A Pilot Randomized Controlled Trial

**DOI:** 10.1101/2025.07.19.25331820

**Authors:** Tomoya Kitade, Takashi Shigematsu, Ichiro Fujishima, Kenjiro Kunieda, Tomohisa Ohno, Satoshi Tanaka

## Abstract

**Background:** Current guidelines recommend combining botulinum toxin type A (BoNT-A) with adjunctive therapies for upper limb spasticity management, but evidence for individualized splinting remains limited and inconsistent.

**Objective:** This pilot randomized controlled trial investigated whether adding individualized splint therapy to BoNT-A enhances therapeutic outcomes compared to BoNT-A alone in chronic stroke patients with upper limb spasticity.

**Methods:** Twenty-six chronic stroke patients with upper limb spasticity were randomized to receive either BoNT-A plus custom-made thermoplastic splint therapy (n=13) or BoNT-A alone (n=13). Both groups received standardized self-training instructions. Primary outcomes were passive range of motion (ROM) for wrist dorsiflexion and middle finger extension. Secondary outcomes included Modified Ashworth Scale (MAS) scores and Disability Assessment Scale (DAS) scores. Assessments were conducted at six time points over 3 months using mixed-effects models for analysis.

**Results:** Both groups demonstrated substantial improvements in wrist dorsiflexion ROM (partial η^2^ = 0.455) and middle finger extension ROM (partial η^2^ = 0.306), significant reductions in MAS scores and improvements in DAS scores. No significant group effects or group × time interactions were observed for ROM, MAS, or DAS measures, indicating equivalent treatment responses.

**Conclusions:** Adding individualized splint therapy to BoNT-A did not provide significant additional benefits for managing upper limb spasticity in chronic stroke patients. Both treatment approaches achieved similar improvements in plasticity and functional outcomes. These findings suggest that for patients who can engage in regular self-training, the addition of a static splint may not offer significant clinical advantages over a 3-month period.

## 1. Introduction

Spasticity, a common complication affecting up to 43% of stroke survivors, significantly impedes upper limb function and compromises rehabilitation potential (1). While botulinum toxin type A (BoNT-A) is recognized as a first-line treatment with Level A evidence for reducing focal spasticity (2), its therapeutic effects are transient, typically lasting 3–4 months. Multiple meta-analyses have demonstrated BoNT-A’s robust efficacy, with effect sizes reaching -0.98 for muscle tone reduction at 4 weeks post-injection (3). Consequently, current practice guidelines strongly recommend combining BoNT-A injections with adjunctive rehabilitation interventions to sustain and optimize clinical outcomes (4).

For lower limbs, Farag et al. demonstrated that casting after BoNT-A improved outcomes compared to other adjunctive therapies (5). However, evidence for upper limb splinting in adult stroke patients remains limited and inconsistent. Santamato et al. found adhesive taping superior to splinting with manual stretching after BoNT-A, but their follow-up was limited to one month (6). Amini et al. reported mixed results when combining BoNT-A with splinting, with some measures of spasticity and function showing improvement while others did not reach statistical significance (7). Recently, Lannin et al. found that adding intensive rehabilitation (including two weeks of casting followed by movement training) to BoNT-A did not improve long-term upper limb function compared to BoNT-A alone (8). These findings highlight a critical knowledge gap and suggest that the effectiveness of adjunctive splinting may depend on the specific intervention protocol and outcome measures used.

A key limitation of previous research is the use of standardized, non-adjustable splints (7) and a primary focus on functional outcomes rather than the direct management of spasticity and ROM (8). This study prioritized spasticity-related outcomes (ROM and MAS) over functional measures to capture the direct mechanical effects of splint therapy. Custom thermoplastic splints provide passive mechanical positioning and sustained stretching, with the most immediate therapeutic impact expected on joint mobility and muscle tone rather than active motor function. This approach allows for more sensitive detection of splint-specific benefits, as functional outcomes reflect multiple contributing factors that may obscure the isolated contribution of adjunctive splinting. The potential benefit of a more personalized approach, using custom-made thermoplastic splints, has been inadequately investigated. This pilot randomized controlled trial (RCT) was therefore designed to address this gap. We hypothesized that adding individualized, custom-made splint therapy to a standard BoNT-A injection protocol would enhance therapeutic outcomes compared to BoNT-A alone.

## 2. Methods

### Study Design

This single-center trial was conducted at Hamamatsu City Rehabilitation Hospital, Japan, between February 2017 and October 2022. Informed consent was obtained from all participants before starting the experiment, which were approved by the Ethics Committee of the Hamamatsu Rehabilitation Hospital (protocol approval no. 16–51, approval date September, 2016). This study was pre-registered in the UMIN Clinical Trials Registry (UMIN-CTR) with the registration number UMIN000032170 (https://center6.umin.ac.jp/cgi-open-bin/ctr/ctr_view.cgi?recptno=R000036702). No significant changes were made to the trial protocol after commencement. Patients were not involved in the design, conduct, or reporting of this trial.

### Participants

Chronic stroke patients (>6 months post-stroke) with upper limb spasticity were enrolled. Based on Julious’s recommendation of a minimum sample size of 12 participants per group for pilot RCT studies (9), we aimed to recruit 15 participants per group (accounting for potential 20% drop-out). Inclusion criteria were: (1) Brunnstrom recovery stage II-III with upper limb spasticity (10); (2) ability to manage splint application (with/without assistance); and (3) no BoNT-A in the previous 12 months. Exclusion criteria included bilateral hemiplegia, severe joint contractures, and BoNT-A contraindications.

### Randomization

Participants were randomly allocated in a 1:1 ratio to either the BoNT-A + splint group or the BoNT-A alone group using a sealed envelope method. The randomization sequence was generated by the first author. Sequentially numbered, opaque, sealed envelopes containing the group assignments were used to ensure allocation concealment. Following enrollment by a treating physician, who was blinded to the randomization sequence, each participant selected an envelope, which was then opened to reveal their group allocation.

### Intervention

Botox® (GlaxoSmithKline) was administered by certified physicians, with injection sites and doses individualized based on clinical assessment using a diagnostic neuromuscular electrical stimulator. Both groups were instructed to perform self-training based primarily on stretching exercises throughout the 3-months post-injection period. Self-training instructions were provided using a standardized checklist form. Occupational therapists prescribed a regimen of 5 sets per day (approximately 10 minutes per set) and verified adherence at each assessment point using a visual analog scale (VAS) ranging from 0% (no adherence to prescribed regimen) to 100% (complete adherence to prescribed regimen).

In the BoNT-A + splint group, an occupational therapist created custom thermoplastic splints (Aquaplast, Sakai Medical Co., Ltd) immediately after injection (Figure 1). The forearm-based volar splints positioned the wrist in dorsiflexion and fingers in extension to the maximum tolerable extent, with particular attention to maintaining the proximal interphalangeal (PIP) and distal interphalangeal (DIP) joints in extension while allowing slight metacarpophalangeal (MP) joint flexion for patient comfort and compliance. Participants were primarily instructed to wear splints during nighttime sleep. For participants who experienced discomfort or sleep disturbance with nighttime wear, daytime use was recommended during periods of inactivity (e.g., during transportation, watching television, or resting periods). Checks and adjustments of splint fit were made during each follow-up outpatient visit. To control for potential variability in treatment effect due to differences in fabrication technique, all splints were made by the same occupational therapist throughout the study.

**Figure 1.**
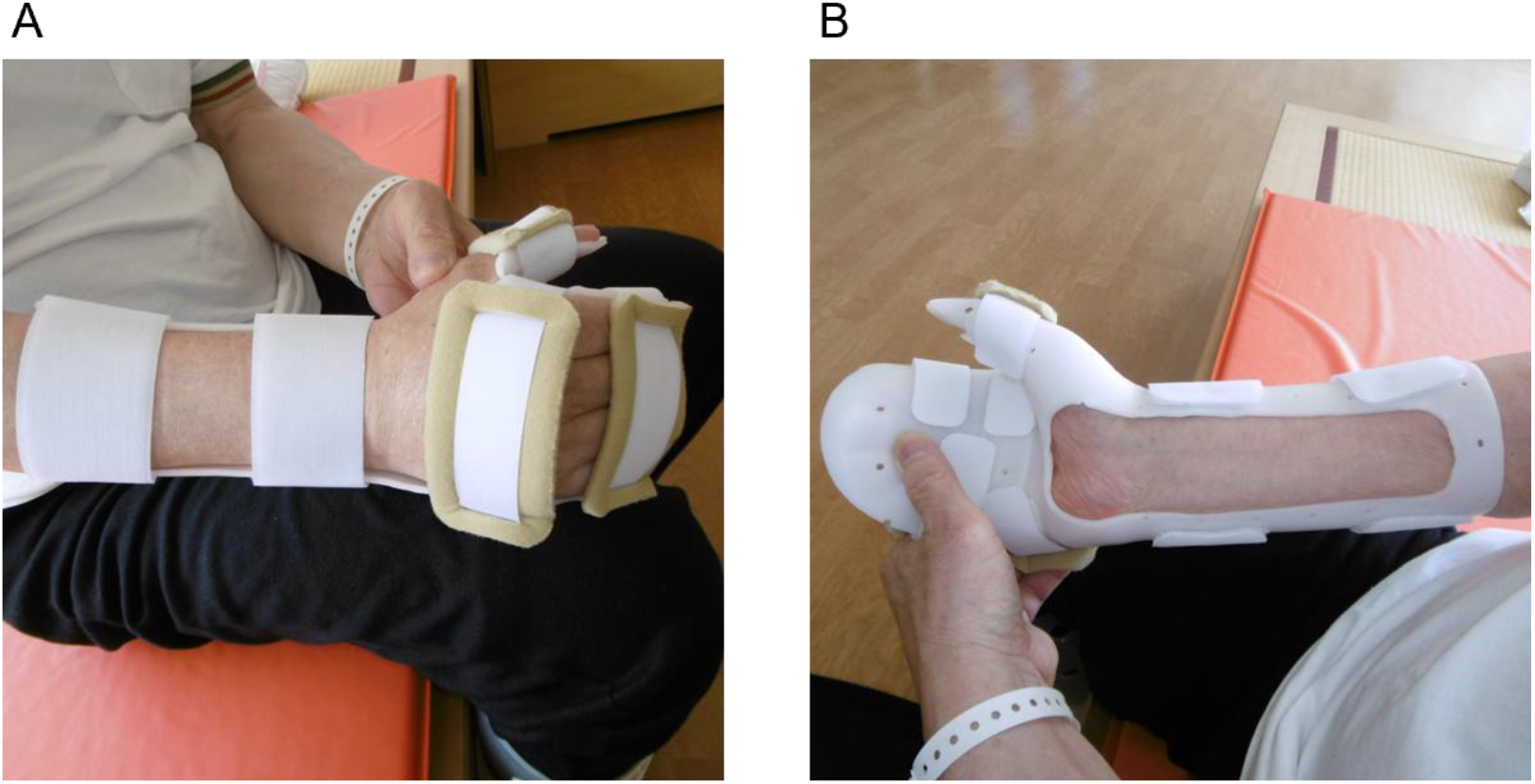
The custom-made thermoplastic splint used for the BoNT-A + splint group. (A) Superior view of the splint as worn. (B) Medial view of the splint. This forearm-based volar splint positioned the wrist in dorsiflexion and the fingers in extension to the maximum tolerable extent.

Adverse events were systematically assessed and documented during regular physician consultations and occupational therapy evaluations throughout the study period. Events of particular interest included splint-related complications such as skin irritation, pressure sores, and increased pain or discomfort.

### Outcome Measures

Although measurements were taken for ROM in various joints (shoulder, elbow, forearm, wrist, fingers, and thumb), our analysis focused specifically on the outcomes most directly affected by the splint intervention. The primary outcome was passive ROM for wrist dorsiflexion and middle finger extension. The middle finger extension angle was calculated as the sum of the passive extension angles of the MP joint, PIP joint, and DIP joint of the middle finger. Middle finger extension was selected because flexor digitorum superficialis (FDS) and flexor digitorum profundus (FDP) muscles originate from single muscle bellies that branch to all fingers, making middle finger measurement representative of overall treatment effects. The splint was designed to maintain these joints in extended position (Figure 1). Secondary outcomes included MAS (11,12) for wrist flexors, FDS, FDP, and flexor pollicis longus, and Disability Assessment Scale (DAS) (13) for hand hygiene, dressing, limb position abnormality, and pain. Assessments were conducted 2-weeks pre-injection, immediately pre-injection, 2 weeks post-injection, and at 1-, 2-, and 3-months post-injection. All outcome measurements were performed by occupational therapists who were not involved in the treatment administration and were blinded to group allocation.

### Statistical Analysis

Mixed-effects models were used to analyze repeated measures data, with time as a within-subjects factor and group as a between-subjects factor. For ROM, MAS and DAS data, linear mixed-effects models were applied. Main effects of time, group, and group × time interactions were examined. Missing data were handled using maximum likelihood estimation within the mixed-effects framework. Effect sizes were reported as partial eta-squared (η^2^) values, with 0.01, 0.06, and 0.14 considered small, medium, and large effects, respectively. Significance was set at p<0.05. All analyses were conducted using R version 4.5.1.

### Reporting Guidelines

This manuscript adheres to the CONSORT 2025 guidelines for reporting RCT (14). A completed CONSORT 2025 checklist is provided as Supplementary Table 1.

## 3. Results

Figure 2 presents the flow diagram. Of 29 patients approached for study participation, 26 were enrolled between February 2017 and October 2022. Twenty-six patients were randomized to either the BoNT-A + splint group (n=13) or BoNT-A alone group (n=13). No significant adverse events occurred during the study. All participants completed the study protocol with no dropouts. One participant in the control group (ID 16) had missing baseline data at the 2-weeks-before time point due to scheduling difficulties. No significant differences were observed in baseline characteristics or botulinum toxin injection parameters (all p > 0.05, Table 1). The median total botulinum toxin dose was 240 units in both groups, with a median of 9 muscles injected per patient. The most frequently targeted muscles were FDS (96% of patients), Brachialis (96%), FDP (92%), biceps brachii (92%), and flexor carpi ulnaris (88%). Adherence to self-training exercises at 3 months post-injection was 62% (interquartile range (IQR): 43-82%) overall, with no significant difference between groups (BoNT-A + splint group: 67%, BoNT-A alone group: 52%, p = 0.14). Self-reported splint wearing time was a mean of 6 hours per day.

**Table 1.**
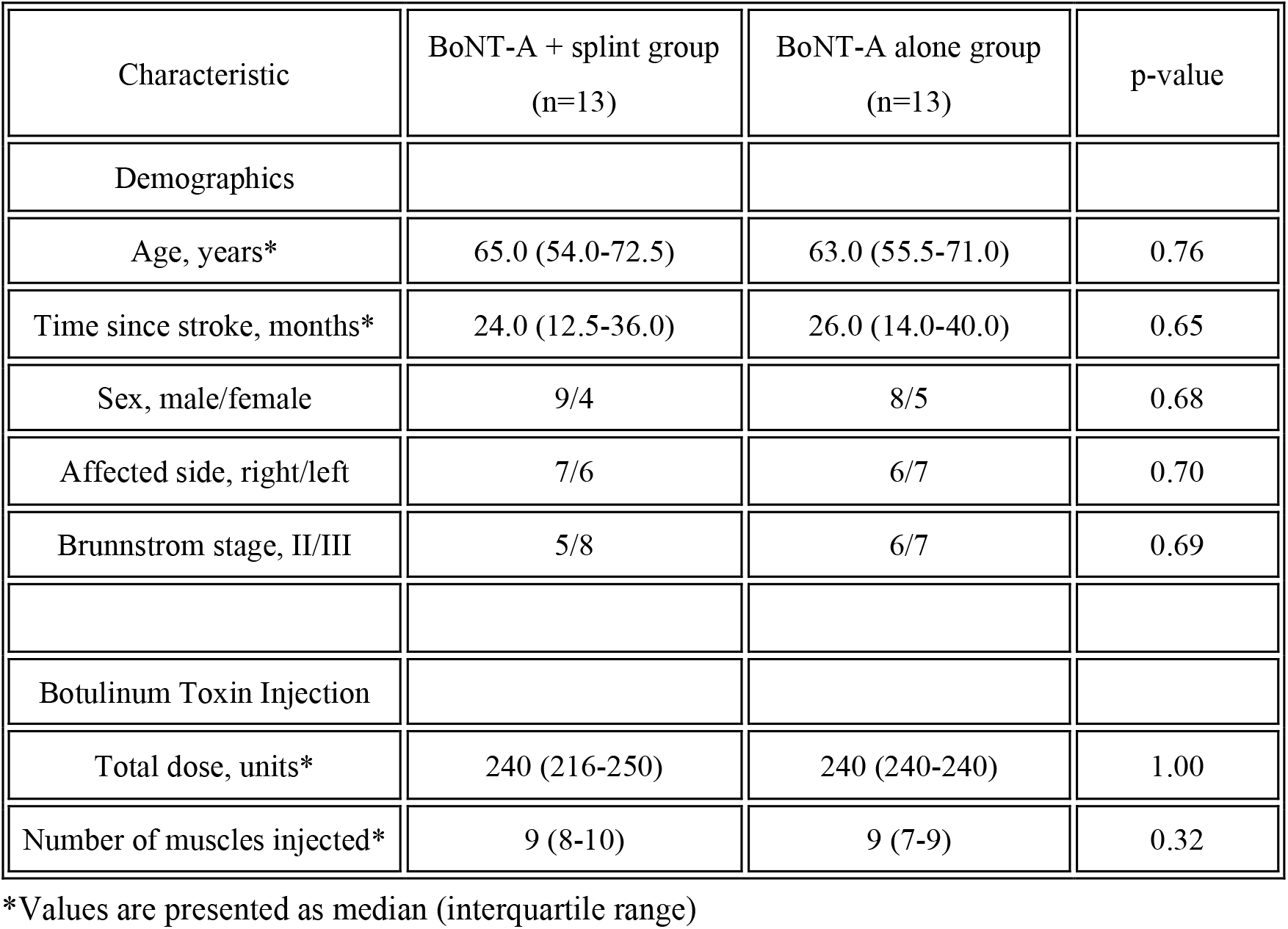
Baseline Characteristics of Participants.

| Characteristic | BoNT-A + splint group<br>(n=13) | BoNT-A alone group<br>(n=13) | p-value |
| --- | --- | --- | --- |
| Demographics |  |  |  |
| Age, years* | 65.0 (54.0-72.5) | 63.0 (55.5-71.0) | 0.76 |
| Time since stroke, months* | 24.0 (12.5-36.0) | 26.0 (14.0-40.0) | 0.65 |
| Sex, male/female | 9/4 | 8/5 | 0.68 |
| Affected side, right/left | 7/6 | 6/7 | 0.70 |
| Brunnstrom stage, II/III | 5/8 | 6/7 | 0.69 |
| Botulinum Toxin Injection |  |  |  |
| Total dose, units* | 240 (216-250) | 240 (240-240) | 1.00 |
| Number of muscles injected* | 9 (8-10) | 9 (7-9) | 0.32 |

\*Values are presented as median (interquartile range)

**Figure 2.**
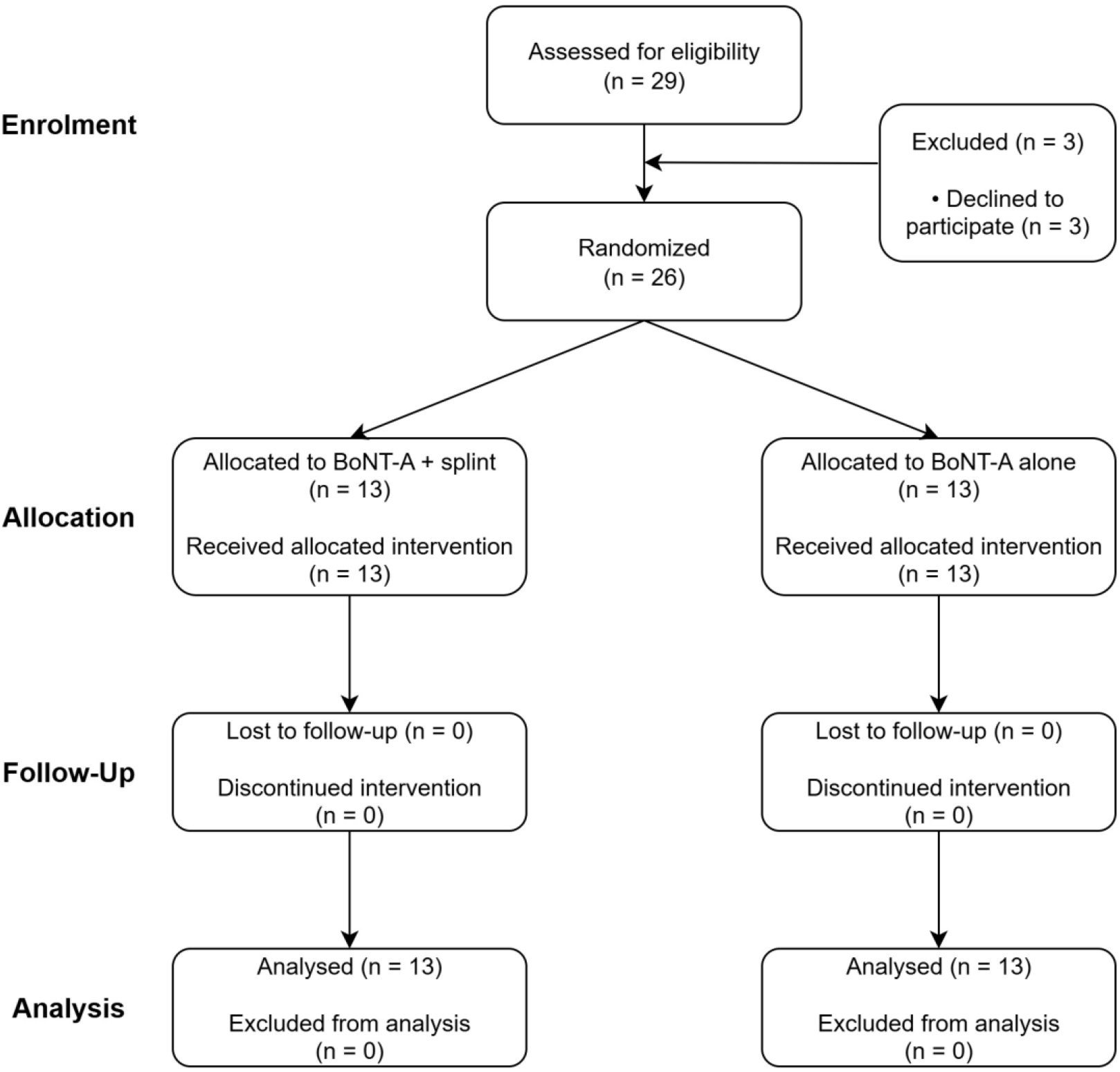
CONSORT flow diagram

### Primary Outcome

Both groups demonstrated improvement in wrist dorsiflexion ROM throughout the study period (Figure 3A). The BoNT-A + splint group showed median values of 50.0^°^ (IQR: 25.0-60.0^°^) at pre-injection, with notable improvement to 60.0^°^ (40.0-70.0^°^ ) at 2 weeks post-injection, and maintained this improvement at 60.0^°^ (45.0-65.0°) at 3 months. The BoNT-A alone group demonstrated a similar trajectory with median values of 40.0^°^ (20.0-50.0^°^) at pre-injection, improving to 55.0^°^ (40.0-60.0^°^) at 2 weeks, and 45.0^°^ (40.0-65.0^°^) at 3 months. Mixed-effects modeling confirmed a significant main effect of time (F(5,119.0) = 19.899, p < 0.001, partial η^2^ = 0.455), indicating a large effect size for temporal changes across both groups. However, no significant group effect was observed (F(1,24.0) = 0.499, p = 0.487, partial η^2^ = 0.020), and the group × time interaction was not significant (F(5,119.0) = 0.710, p = 0.617, partial η^2^ = 0.029). These findings indicate that while both groups experienced substantial improvement in wrist dorsiflexion ROM following BoNT-A injection, the addition of splint therapy did not provide significant additional benefit over the 3-month observation period.

**Figure 3.**
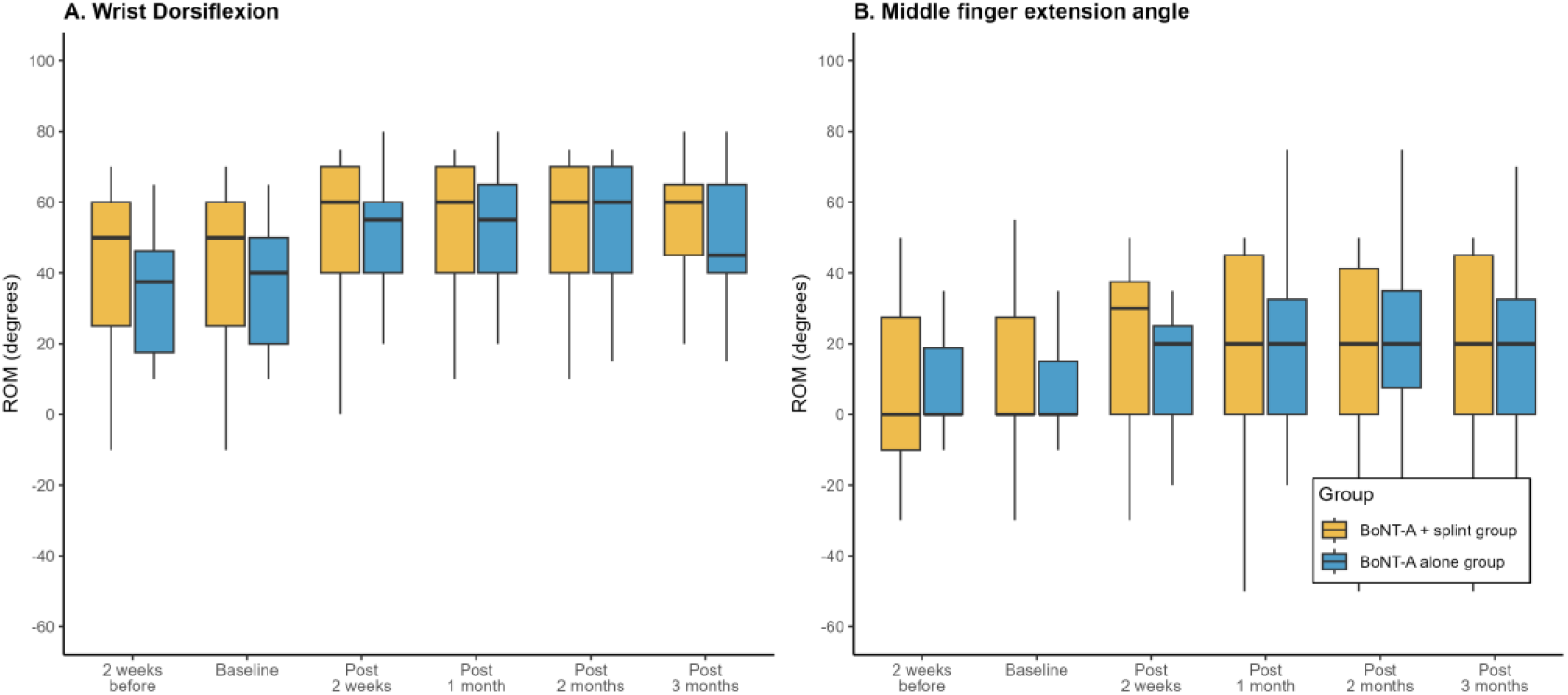
Changes in ROM over time following botulinum toxin injection. (A) Wrist dorsiflexion ROM and (B) Middle finger extension angle measured at six time points: 2 weeks before injection, baseline, and 2 weeks, 1 month, 2 months, and 3 months post-injection. Data are presented for BoNT-A + splint group (orange) and BoNT-A alone group (blue). Box plots show median (center line), interquartile range (Q1-Q3; box), and whiskers extending to the furthest data points within 1.5× interquartile range from quartiles.

For middle finger extension, both groups showed progressive improvement from baseline to 3 months post-injection (Figure 3B). The BoNT-A + splint group progressed from a median of 0.0° (−30.0 to 10.0^°^) at pre-injection to 20.0^°^ (0.0 to 30.0^°^) at 3 months. Similarly, the BoNT-A alone group improved from 0.0° (−10.0 to 0.0°) at pre-injection to 20.0^°^ (0.0 to 30.0^°^) at 3 months, demonstrating comparable improvement patterns between groups. Mixed-effects analysis revealed a significant main effect of time (F(5,119.0) = 10.474, p < 0.001, partial η^2^ = 0.306), representing a large effect size for temporal improvement. No significant group effect was detected (F(1,24.0) = 0.101, p = 0.753, partial η^2^ = 0.004), and the group × time interaction was not significant (F(5,119.0) = 0.798, p = 0.553, partial η^2^ = 0.032). These results demonstrate that both treatment approaches achieved similar gains in finger extension ROM, with no additional benefit from adjunctive splint therapy over BoNT-A alone.

### Secondary Outcomes

For MAS, all muscle groups demonstrated substantial spasticity reduction over the 3-month period in both treatment groups (Figure 4). Mixed-effects modeling confirmed significant temporal improvements across all four muscle groups, with effect sizes ranging from medium to large. However, no significant group effects or group × time interactions were detected for any MAS measure (all p > 0.05), indicating equivalent spasticity reduction between treatment approaches.

**Figure 4.**
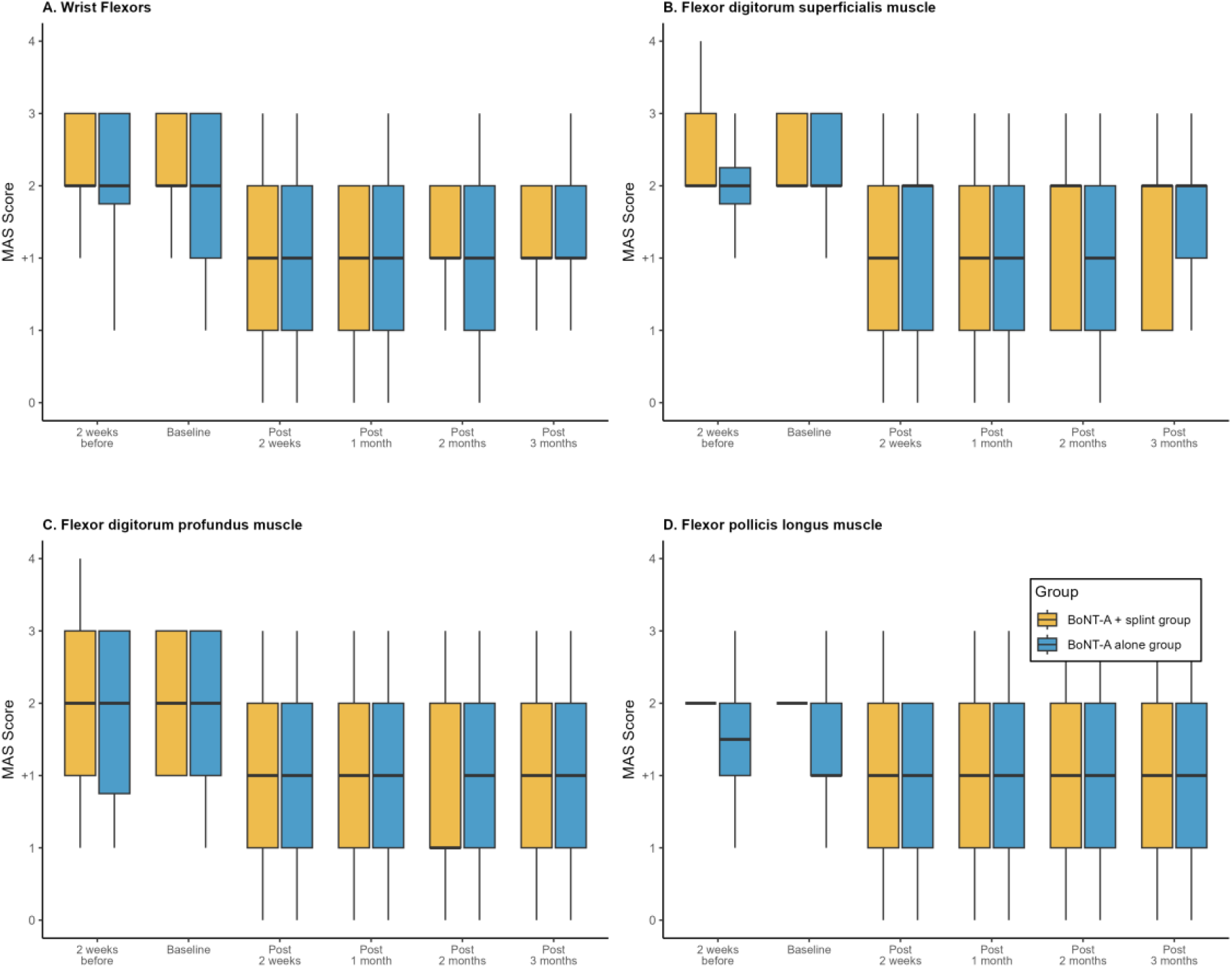
Changes in MAS scores over time following botulinum toxin injection. (A)Wrist Flexors, (B) Flexor digitorum superficialis muscle, (C) Flexor digitorum profundus muscle, and (D) Flexor pollicis longus muscle measured at six time points: 2 weeks before injection, baseline, and 2 weeks, 1 month, 2 months, and 3 months post-injection. Data are presented for BoNT-A + splint group (orange) and BoNT-A alone group (blue). Box plots show median (center line), interquartile range (Q1-Q3; box), and whiskers extending to the furthest data points within 1.5× interquartile range from quartiles

Both groups demonstrated meaningful functional improvements across all DAS domains over the 3-month study period (Figure 5). Mixed-effects analysis revealed significant temporal improvements across all four DAS domains, with large effect sizes confirming substantial functional benefits. No significant group effects or group × time interactions were detected for any DAS measure (all p > 0.05), although pain showed medium between-group effect sizes without reaching statistical significance (partial η^2^ = 0.098, p=0.12). These results indicate that both treatment approaches achieved similar functional improvements.

**Figure 5.**
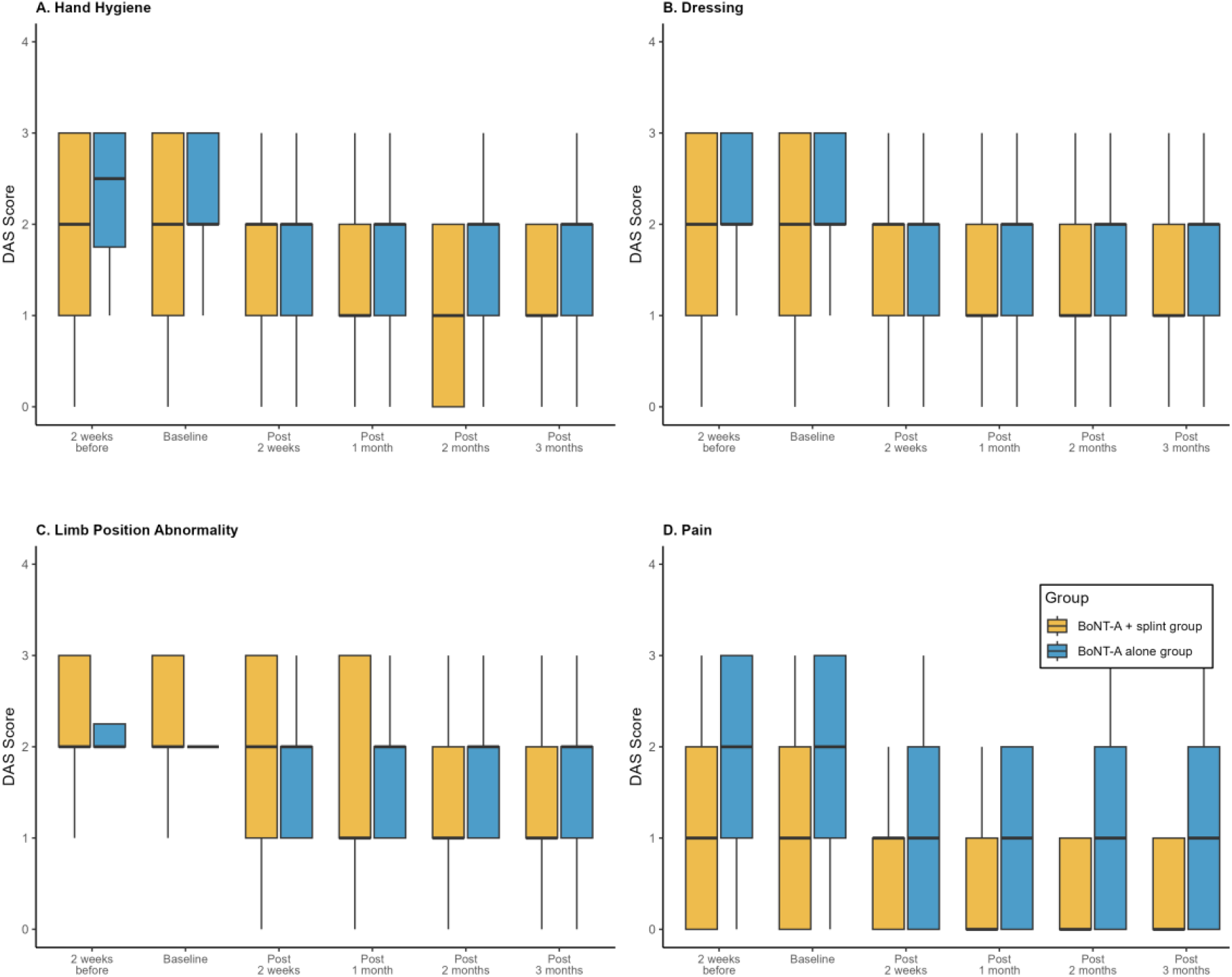
Changes in DAS scores over time following botulinum toxin injection. (A) Hand Hygiene, (B) Dressing, (C) Limb Position Abnormality, and (D) Pain measured at six time points: 2 weeks before injection, baseline, and 2 weeks, 1 month, 2 months, and 3 months post-injection. Data are presented for BoNT-A + splint group (orange) and BoNT-A alone group (blue). Box plots show median (center line), interquartile range (Q1-Q3; box).

## 4. Discussion

This pilot study found no significant additional benefit of individualized splint therapy when combined with BoNT-A for upper limb spasticity management in chronic stroke patients. Both groups showed similar improvements in ROM, MAS, and DAS following BoNT-A injection.

Our findings align with those of Santamato et al. (6), who also reported no significant additive effects of BoNT-A plus standardized splinting compared to other interventions. Our study is distinctive, however, in its use of individually tailored thermoplastic splints, a methodological enhancement intended to optimize the therapeutic effect. Despite this personalized approach, the lack of significant between-group differences suggests that the primary driver of improvement in our study may have been the potent effect of BoNT-A combined with self-training, rather than the passive mechanical positioning offered by the splint. These results are also consistent with Lannin et al.’s finding that intensive rehabilitation (including casting) following BoNT-A provided no additional benefit compared to BoNT-A plus usual care (8). However, important methodological differences exist. While Lannin’s study used only two weeks of casting followed by exercise, our protocol maintained splinting throughout the 3-month period. Additionally, Lannin focused primarily on functional improvement, whereas our study specifically targeted spasticity.

Two main factors may explain the lack of significant between-group differences. First, the potent therapeutic effect of BoNT-A, evidenced by the large within-group effect sizes for ROM improvement (e.g., partial η^2^=0.455 for wrist dorsiflexion), may have created a ceiling effect, masking any smaller, additive benefits of splinting. Meta-analyses have consistently demonstrated that BoNT-A produces large effect sizes for spasticity reduction, with standardized mean differences reaching -0.98 for muscle tone at 4 weeks post-injection (3). Importantly, systematic reviews indicate that BoNT-A has favorable effects on passive ROM measures, but limited effects on active functional use of the arm and hand (15). Since our primary outcomes were passive ROM measures, this “ceiling effect” hypothesis is supported by existing evidence.

Second, the self-training protocol appears to have been a powerful intervention. Maintaining adherence to home-based exercise is a challenge across chronic diseases, with non-adherence rates often exceeding 50% (16). In this context, the overall self-reported adherence of 62% in our cohort can be considered moderate. Indeed, a scoping review on post-stroke home programs revealed that achieving adherence rates of 75% or higher is uncommon (17). It is plausible that this moderate level of patient engagement provided a therapeutically sufficient dose of active exercise, which, combined with the potent effects of BoNT-A, drove the significant improvements observed in both groups. In fact, research has demonstrated that BoNT-A combined with home-based exercise programs can produce sustained motor improvements (18). This may have in turn created a ceiling effect, masking any smaller, additive benefits of passive splinting. Future research should investigate whether implementing strategies to enhance motivation for self-training participation could further optimize therapeutic outcomes in post-stroke spasticity management (19-23).

Despite the lack of significant between-group differences, several clinical insights emerge from this study. Both groups demonstrated clinically meaningful improvements in ROM and MAS, confirming the effectiveness of BoNT-A as a cornerstone treatment for upper limb spasticity. The large effect sizes observed for temporal changes indicate that BoNT-A alone provides substantial therapeutic benefit. The absence of additional benefit from individualized splinting may reflect the complexity of post-stroke spasticity management, where multiple factors beyond mechanical positioning influence recovery. Our findings suggest that in motivated patients who can perform regular self-training, the addition of splinting may not provide meaningful clinical advantages within the 3-month timeframe. In addition, while not statistically significant, the medium between-group effect sizes observed for pain on the DAS might suggest a potential clinical trend. This trend aligns with evidence that BoNT-A provides significant pain reduction in post-stroke spasticity (24). A larger sample size might be needed to detect potential additive pain relief benefits of combining BoNT-A with custom-made splint therapy.

Several limitations must be acknowledged. First, as a pilot study, the small sample size (n=26) limited our statistical power to detect small but potentially meaningful between-group differences. Second, self-report measures might be susceptible to overestimation due to recall and social desirability biases (25). Therefore, the 62% adherence rate could be interpreted as a potential upper estimate of actual performance. Third, the 3-month follow-up period may have been too short to evaluate the long-term effects of splinting on contracture prevention. Finally, the study population was limited to chronic stroke patients with Brunnstrom stage II-III spasticity, and the findings may not be generalizable to patients with different severity levels or in different stages of recovery.

## 5. Conclusion

Adding individualized splint therapy to BoNT-A did not provide significant additional benefits for managing upper limb spasticity in chronic stroke patients over a 3-month period. Both treatment groups demonstrated substantial improvements in spasticity and functional outcomes, suggesting that BoNT-A therapy combined with self-training exercises is a highly effective treatment approach. Future studies with longer follow-up periods and stratification of patients based on clinical characteristics are needed to further explore the potential role of adjunctive therapies in post-stroke spasticity management.

## Acknowledgments

The authors utilized an AI-assisted tool (Claude 4.0 Sonnet by Anthropic) for English editing. All content was thoroughly reviewed and verified by the authors before submission.

## Supplementary Materials

Supplementary Table 1: CONSORT 2025 Checklist

## Data Availability Statement

Data are available upon reasonable request to the corresponding author.

## Author Contributions

Study concept and design: T. Kitade, I. Fujishima, T. Shigematsu, K. Kunieda, T. Ohno

Data acquisition: T. Kitade, I. Fujishima, T. Shigematsu

Data analysis: T. Kitade, S. Tanaka

Drafting of the manuscript: T. Kitade, S. Tanaka

Critical revision of the manuscript: All authors

Study supervision: I. Fujishima, T. Shigematsu

